# Digital Twin Model of Treatment Outcomes in Post-Stroke Aphasia

**DOI:** 10.64898/2026.02.03.26345022

**Authors:** Natalie Busby, Nicholas Riccardi, Janina Wilmskoetter, Eleanor Jeakle, Roger Newman-Norlund, Sigfus Kristinsson, Chris Rorden, Julius Fridriksson, Leonardo Bonilha

## Abstract

**Background:** Recovery from chronic post-stroke aphasia is highly heterogeneous and shaped by lesion characteristics, brain integrity, and systemic health. Traditional group-level models struggle to capture this multidimensional, dynamic variability. Digital twin approaches - patient-specific, continually updating models - may enable individualized prediction and counterfactual evaluation of modifiable risk factors. Therefore, the aim was to develop and validate a proof-of-concept digital twin that predicts individual naming outcomes during language treatment and quantifies the estimated impact of modifiable health factors on naming. This study represents the first application of digital twin modeling to aphasia recovery, and we hypothesize that this could constitute a critical first step toward dynamically adaptive, personalized models for aphasia rehabilitation.

**Methods:** We analyzed longitudinal data from 106 chronic stroke survivors with aphasia enrolled in the POLAR randomized clinical trial. For each participant we combined baseline demographic/health variables (age, sex, education, days post-stroke, hypertension, diabetes, BMI), lesion load in left-hemisphere language ROIs (JHU atlas), ROI-level white-matter microstructure (FA), and resting-state functional connectivity restricted to language regions. A continual-learning linear model (River framework; Adam optimizer) was pretrained on baseline data and updated across timepoints. Model performance was assessed by R² at the final timepoint. Counterfactual simulations systematically altered hypertension, diabetes, and BMI to estimate isolated and combined effects on predicted Philadelphia Naming Test (PNT) scores.

**Results:** The digital twin predicted final PNT scores with R² = 0.5848 (explaining approximately 58% of variance). The largest contributors were prior naming performance, age, lesion load in language regions, and white-matter integrity in temporal regions (notably right MTG and STG pole). Counterfactual results estimated modest but consistent effects of health factors, with them collectively accounting for approximately 25% of the variance in treatment gains. The average change in PNT score with counterfactual changes was 7.92 (SD = 16.11). Therefore, diabetic status explained 2% of the variance in treatment gains, hypertensive status explained 4.75%, and increasing BMI explained 18.5%.

**Conclusions:** This study demonstrate the feasibility and clinical potential of applying a digital twin framework to chronic post-stroke aphasia, with the model successfully predicting more than half the variance in naming performance during language treatment. Through counterfactual simulation, we demonstrated that modifiable health factors exert measurable, bidirectional influences on predicted treatment outcomes, underscoring the role of systemic health in shaping language recovery. Although the individual effects of these factors were modest in magnitude, their cumulative influence on treatment gains illustrates how multiple small biological contributors can add up to shape meaningful differences in language outcomes. More broadly, these findings illustrate the potential value of digital twin models for aphasia treatment, particularly as a tool to integrate diverse biological factors and generate individualized, dynamically updated predictions.

**Graphical Abstract:** 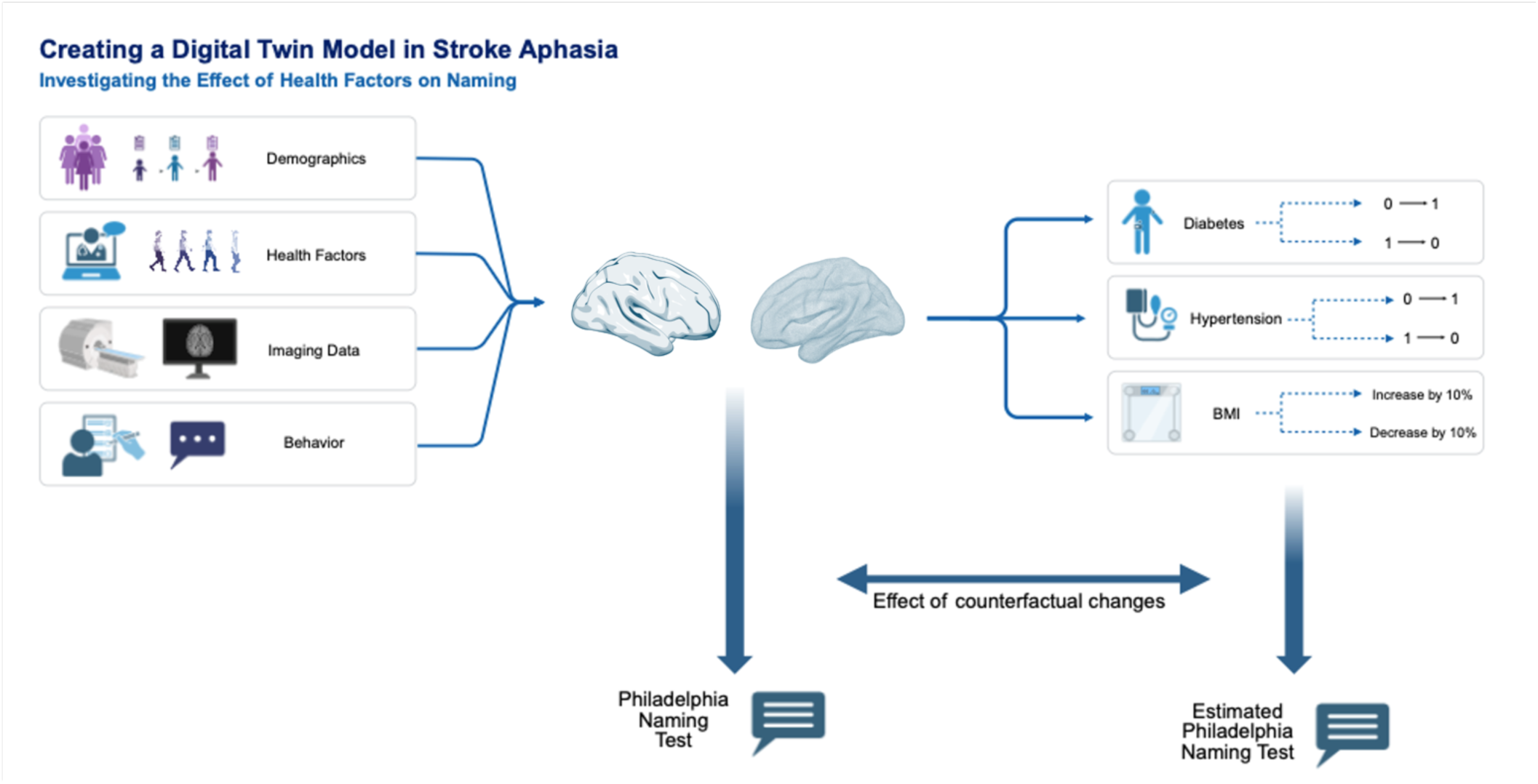

## Introduction

Speech therapy can lead to language improvements for many individuals with chronic aphasia even after many years beyond the original stroke^1,2^. Nonetheless, treatment response is highly variable. While some individuals can achieve significant gains, others do not experience meaningful changes. Unfortunately, the mechanisms underlying treatment response, aphasia recovery (and neurological rehabilitation in general) are not well-understood. Naturally, stroke lesion factors such as size and location play a significant role in leading to the development of aphasic symptoms and to their presence in the chronic stages. However, lesion factors alone contribute to only a proportion of the recovery variance^3–6^. For this reason, the long-term recovery and response to treatment interventions are critically influenced by the interaction between lesion factors and other neurobiological factors that promote plasticity and recovery.

Moreover, neurobiological recovery is a dynamic, time-dependent process shaped by the progressive capacity of neural tissue to reorganize in response to therapeutic stimuli. This is particularly relevant for interventions that actively engage neuroplastic mechanisms. Despite the acknowledged significance of dynamic neurobiological properties, there is a notable scarcity of methods for quantitatively evaluating how evolving neuroplasticity influences individual treatment response trajectories.

More recent studies have confirmed that aphasia recovery depends on health factors that contribute to the integrity of the neurological tissue beyond the lesion, i.e., the residual brain tissue after stroke. Moreover, demographic and medical risk factors, such as age or cardiovascular status^7–12^ are becoming increasingly recognized as key contributors to overall brain health and cognition among older adults^13–19^, and emerging evidence indicates that these same factors substantially shape language recovery in aphasia^2,19–23^. This is a promising new path for a mechanistic and dynamic understanding of aphasia outcomes and clinical outcome prediction.

Nonetheless, aphasia recovery studies are complex due to variability not only in lesion location but also in medical and cardiovascular factors that could influence recovery potential. This multifactorial variability can pose challenges for statistical modeling, particularly in the context of aphasia study cohorts with small sample sizes. Predicting language outcomes is thus inherently complex due to numerous individual factors that likely exert influence such as demographic variables (e.g., age, education), vascular and metabolic health markers (e.g., hypertension, diabetes, BMI), indices of brain integrity (e.g., lesion volume, white matter microstructure), and measures of functional network reorganization (e.g., resting-state fMRI connectivity patterns). Moreover, these factors can interact non-linearly. Therefore, traditional group-level analytical approaches can be limited in their ability to adequately capture the complex, multidimensional interactions among contributing factors.

These challenges underscore the need for individualized prediction models capable of integrating complex, multidimensional data dynamically over time. Digital twin models provide a computationally efficient framework for simulating the interaction between contributing factors at the individual level, rather than relying on population-averaged effects that may obscure clinically meaningful individual variability. By modeling how lesion characteristics, vascular health, brain integrity, and functional reorganization interact dynamically within each person’s unique biological context, digital twins can generate personalized predictions that evolve as new data are acquired throughout the treatment course. This approach offers a promising path toward understanding individual recovery mechanisms.

First proposed by Michael Grieves^24,25^, a digital twin model is a virtual replica of a process, individual, or physical system that allows modelling of the interaction between the modeled system and the environment, i.e., how does the system respond to perturbations and why. The difference between more traditional predictive models and digital twins lies primarily in the idea that a digital twin model is dynamic, and the addition of new data informs, adjusts and shapes the model^26,27^. Digital twin models typically incorporate rich, multidimensional data. Historically, digital twin models have been first used in the military and aerospace research^25^, city planning^28^ and architecture^29^. More recently, in a 2022 review of digital twin modelling in the literature, nearly half of the publications using digital twin modelling were related to manufacturing^25,30^. While this type of modelling is becoming increasingly widespread, it remains an emerging but growing concept in biological settings. Early digital twin studies have been used for diagnostic testing^31^, identification of treatment response^32,^ and identification of tailored treatment options, for example, in cardiac physiology^33,34^.

In neurological rehabilitation, digital twin modelling remains a new concept. In a study by Lauer-Schmaltz and colleagues (2024) a badminton game with a digital twin opponent was developed ^35^ to simulate learning experiences. Zhang et al. also investigated digital twin approaches for data visualization rather as opposed to clinical prediction ^36^.

This study represents the first application of digital twin modeling to aphasia recovery. Although the complexity of interacting neurobiological, vascular, and functional factors ultimately motivates this approach, here, we begin by testing whether fundamental demographic and health variables can be effectively integrated within a digital twin framework to generate dynamic individualized predictions. Our goal is to provide a foundational investigation that establishes proof-of-concept for the methodology while creating a scalable platform that can progressively incorporate more complex multimodal and neuroimaging data as they become available. By validating the approach with accessible baseline factors, we hypothesize that this could constitute a critical first step toward dynamically adaptive, personalized models for aphasia rehabilitation.

## Methods

### Participants

We evaluated a longitudinal prospective cohort of stroke survivors with aphasia (N = 106; 63 males, 43 females) who took part in the randomized clinical trial titled POLAR (<u>P</u>redicting <u>O</u>utcomes of <u>La</u>nguage <u>R</u>ehabilitation)^37^, clinical trial (NCT03416738). Data were collected at research laboratories at the Center for the Study of Aphasia Recovery (C-STAR) at the University of South Carolina (Columbia, SC), and at the Medical University of South Carolina (Charleston, SC). ASHA-certified speech-language pathologists (SLPs) with experience working with individuals with aphasia administered all assessments and treatments. Aphasia type was determined based on the Western Aphasia Battery Revised (WAB-R)^38^ subscores following published norms.

The following inclusion/exclusion criteria were used in the POLAR trial. Participants were included in the study if they i) incurred a left-hemisphere ischemic or hemorrhagic stroke to the middle cerebral artery, ii) had chronic aphasia (≥ 12 months post-stroke), iii) were between 21 and 80 years of age, iv) had to have spoken English as their primary language for at least 20 years, and v) were able to provide written or verbal consent. Participants were excluded if they had i) severely limited verbal output (as measured by a WAB-R Spontaneous Speech rating scale score of 0-1), ii) severely impaired auditory comprehension (as measured by a WAB-R Auditory Comprehension rating scale score of 0-1), iii) bilateral or cerebellar stroke, or iv) or contra-indications to testing with magnetic resonance imaging (MRI). Individuals with multiple strokes were eligible if all lesions were confined to the left supratentorial regions.

Health and demographic data were recorded at the time of recruitment. During this baseline testing, participants were given a detailed questionnaire that addressed their medical history, including yes/no questions about having diabetes and hypertension. BMI was calculated based on their weight and height.

### Behavioral Data

Language assessments were administered and scored by ASHA-certified SLPs at the time of enrollment. One of these assessments was the WAB-R Aphasia Quotient (AQ^95^) where scores range from 0-100 and ≤ 93.8 is indicative of aphasia. The other was the Philadelphia Naming Test (PNT^96^) which was administered twice, and the scores were averaged to serve as the baseline PNT score. After baseline testing, participants completed semantic and phonological therapy with the following timeline: three weeks of treatment were following by 4 weeks of rest and then 3 weeks of treatment (Figure 1). Treatment type was counterbalanced across participants, half received semantic treatment first, and the other half received phonological treatment first. See Kristinsson et al.^37,39^ for the detailed protocol.

**Figure 1.**
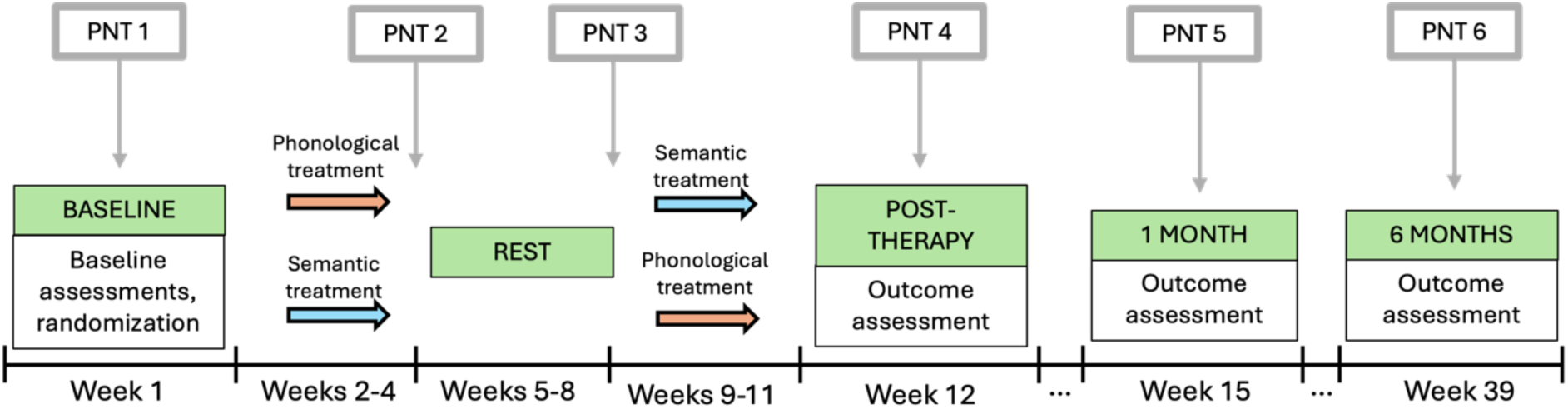
The study design timeline, including when each Philadelphia Naming Test (PNT) was administered.

### MRI Data Acquisition and Preprocessing

#### T1-weighted and T2-weighted Imaging

At enrollment, all participants underwent high-resolution T1- and T2-weighted neuroimaging on a Siemens Trio 3T scanner equipped with a 12-channel (Trio configuration) or 20-channel (following upfit to Prisma configuration) head coil using the following parameters: T1-weighted imaging utilized an MP-RAGE sequence with 1 mm isotropic voxels, a 256 x 256 matrix size, a 9° flip angle, and a 92-slice sequence with repetition time (TR)=2250 ms, inversion time (TI)=925 ms, and echo time (TE)=4.11 ms. T2-weighted scans were acquired using the same angulation and volume center as the T1 scan. This 3D T2-weighted SPACE sequence used a resolution of 1 mm^3^ was used with a field of view=256 x 256mm, 160 sagittal slices, variable degree flip angle, TR=3200 ms, TE=212ms, x2 GRAPPA acceleration (80 reference lines).

#### Lesion Identification

Lesions were drawn onto participants’ T2-weighted image by a neurologist (author LB) or trained study staff member (author RNN), both of whom were blinded to participant demographic information and WAB-R scores at the time of the lesion drawing. The participants’ T2 images were co-registered to their T1 images, and the resulting function was then used to spatially transform the binary lesion maps into native T1 space. Resliced lesion maps were smoothed with a 3mm full-width half maximum Gaussian kernel to remove sharp edges associated with hand drawing. Enantiomorphic segmentation-normalization was then conducted using the *nii_preprocess* pipeline (https://github.com/neurolabusc/nii_preprocess)^40^, a series of custom MATLAB-based (R2017b, TheMathWorks) scripts^40^ that leverage multiple best-of-breed programs (SPM12; Functional Imaging Laboratory, Wellcome Trust Centre for Neuroimaging, Institute of Neurology [www.fil.ion.ucl.ac.uk/spm], FSL v6.0.3^41^ <u>and MRItrix</u> [https://www.mrtrix.org/]) to normalize and process MRI data acquired from participants with lesioned brains. This included creating of a mirrored image of the right hemisphere, which was co-registered to the native T1 image. A chimeric image (i.e., a healed brain) was then created, based on the native T1 scan with the lesioned tissue replaced by tissue from the mirrored hemisphere. SPM12’s unified segmentation-normalization^42^ warped this chimeric image to standard space, and the resulting spatial transform was applied to the native T1 scan as well as the lesion map and the T2/DWI image.

#### DiHusion-weighted imaging

Diffusion tensor imaging was acquired in a pair with b = 1000 s/mm^2^ (43 volumes of which 7 were b = 0, TR = 5,250 ms, TE = 80.0 ms) and a pair with b = 2,000 s/mm^2^ (56 volumes of which 6 were b = 0, TR = 5,470 ms, TE = 85.4 ms, TA = 5:23). The pairs were identical except for reversed phase encoding polarity (A > P vs. P > A). Scans used a monopolar sequence with a 140 × 140 matrix, 210 × 210 mm FOV, multiband ×2, 6/8 partial Fourier, 80 contiguous 1.5-mm axial slices. Images were denoised using Mrtrix3’s dwidenoise function to reduce the thermal noise and Gibbs ringing artifacts. Denoised images were corrected for eddy current distortions, motion and susceptibility-induced off-resonance effects using FSL’s eddy and topup tools.

Following distortion correction, diffusion tensors were fitted voxelwise using FSL’s dtifit, which computes scalar diffusion parameters including fractional anisotropy (FA). The resulting FA images represent the normalized standard deviation of the three eigenvalues of the diffusion tensor, reflecting white matter microstructural integrity. The FA maps were subsequently masked to remove spurious edge artifacts and then spatially normalized to each subject’s brain-extracted T1-weighted anatomical image using SPM12’s affine and nonlinear registration normalization procedure. An FA score for each region of the JHU atlas was extracted from each individual’s normalized FA maps.

#### Resting-state functional MRI Data

The resting-state functional MRI (rs-fMRI) data were acquired using the following imaging parameters: a multiband sequence (2) with a mm field of view, a matrix size, and a 72-degree flip angle, 50 axial slices (2 mm thick with gap yielding 2.4 mm between slice centers), repetition time TR = 1650 ms, TE = 35 ms, GRAPPA = 2, 44 reference lines, interleaved ascending slice order. During the scanning process, the participants were instructed to stay still with eyes closed. A total of 370 volumes were acquired. Data were corrected for motion using the Realign and Unwarp procedure in SPM12 with default settings. Brain extraction was then performed using the SPM12 script pm_brain_mask with default settings. Slice time correction was also done using SPM12. The mean fMRI volume for each participant was then aligned to the corresponding T2-weighted image to compute the spatial transformation between the data and the lesion mask. The fMRI data were then spatially smoothed with a Gaussian kernel with FWHM= 6 mm. To eliminate artifacts driven by lesions, a pipeline proposed by Yourganov et al. (2018) was applied to the rs-fMRI^43^. The FSL MELODIC package was used to decompose the data into independent components (ICs) and to compute the Z-scored spatial maps for the ICs. The spatial maps were thresholded at and compared with the participant’s lesion mask. The Jaccard index, computed as the ratio between the numbers of voxels in the intersection and union, was used to quantify the amount of spatial overlap between the lesion mask and thresholded IC maps, both of which were binary. ICs corresponding to the Jaccard index were deemed significantly overlapping with the lesion mask and then regressed out of the fMRI data using the fsl_regfilt script from the FSL package. By applying the Johns Hopkins atlas, 189 regions of interest (ROIs) were created.

### Preparing the Data

For each participant, we constructed a multimodal feature set consisting of demographic and health-related variables (age, sex, years of education, days post-stroke, hypertension status, diabetes status, and BMI). The primary outcome variable was performance on the Philadelphia Naming Test (PNT). Lesion burden was quantified by calculating the proportion of each language-related ROI in the Johns Hopkins atlas that overlapped with the participant’s lesion. Language-related ROIs included the angular gyrus (AG), inferior frontal gyrus (IFG) triangularis and opercularis, middle frontal gyrus (MFG), middle temporal gyrus (MTG), posterior superior temporal gyrus (PSTG), posterior supramarginal gyrus (PSMG), superior temporal gyrus (STG), and the STG pole in the left hemisphere.

Similar to the lesion data, to incorporate structural and functional integrity without inflating model dimensionality, we used only language-related ROIs. For diffusion MRI, fractional anisotropy (FA) values were extracted for the left and right AG, IFG triangularis and opercularis, MFG, MTG, PSTG, PSMG, STG and the STG pole. An analogous approach was applied to resting-state fMRI: ROI-level connectivity values This strategy preserved sensitivity to hemispheric differences significant for capturing changes in the stroke-affected hemisphere, while reducing the risk of overfitting that could arise from including numerous region-specific diffusion and connectivity measures as a single, high-dimensional vector. Missing values were imputed using mean imputation and extreme values were constrained to the 1^st^-99^th^ percentile to reduce the influence of outliers. Skewed predictors were log transformed (log1p-transformed), numeric features were standardized using StandardScaler and categorical variables underwent one-hot encoding. See Figure 2 for a flow chart of the main methods.

**Figure 2.**
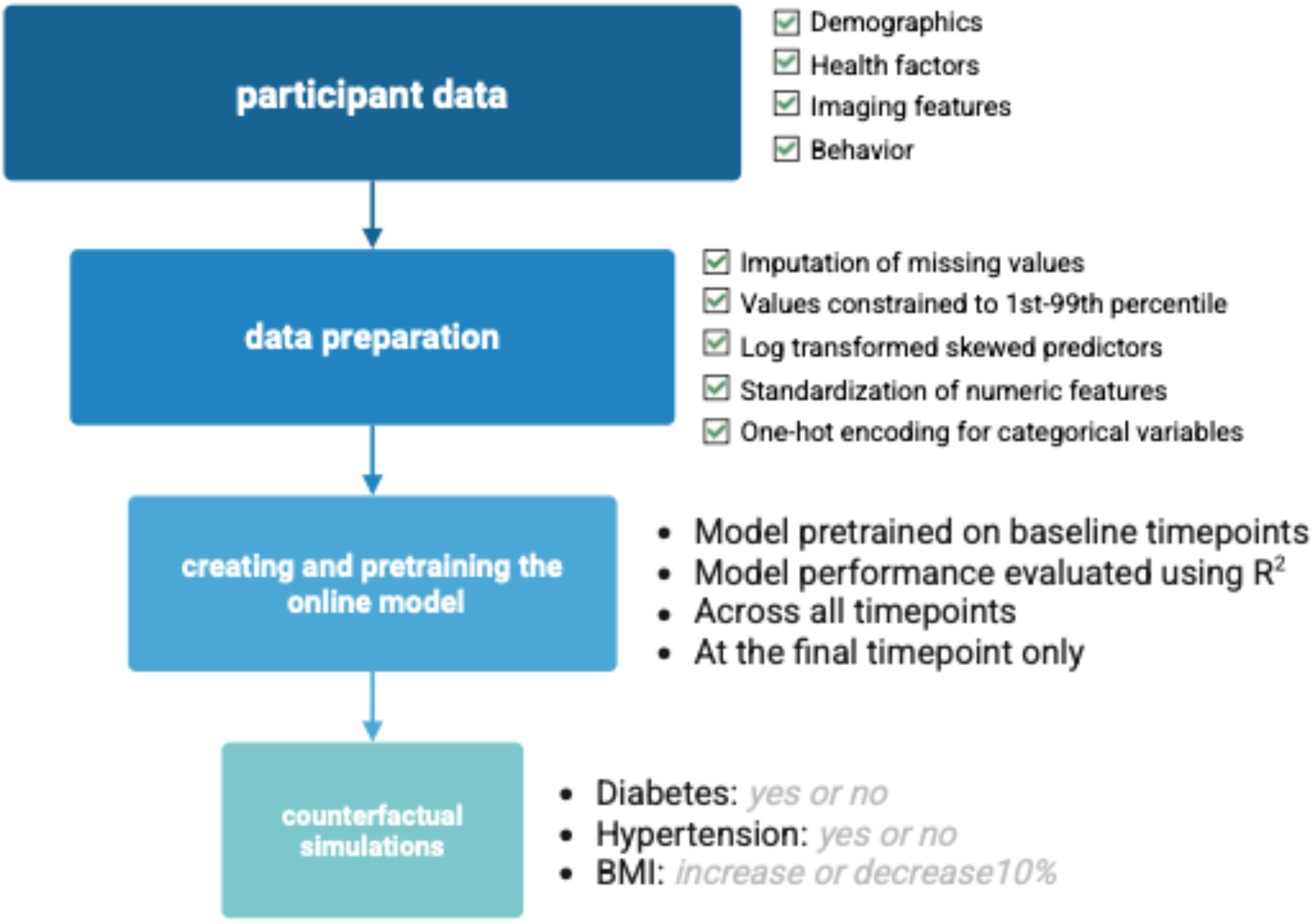
Flow chart to outline how the model was set up.

### Modeling Approach

We used a continual learning framework implemented with the River Python package, which allows for online incremental model updates with each new observation. The regularized linear regression model was trained with the adaptive movement estimation (Adam) optimizer which updates weights gradually based on streaming data. The learning rate was set to 0.0001 to ensure careful weight adjustments, given that it was an online model. Model performance was evaluated using R^2^ at the final timepoint.

To improve model stability, we pretrained the model on the baseline data before performing online learning on the subsequent timepoints (Figure 2). This mirrors how a real-time digital twin model would learn from the first timepoint before additional observations were collected.

### Counterfactual Simulations

Using the trained model, we simulated counterfactual scenarios for each participant through systematically modifying the health variables while keeping all other features constant. Predictions were recomputed after each ‘change’ to estimate the isolated effect of that variable. Counterfactual changes included changing the hypertension status from 0 to 1, and vice versa. Similarly, diabetes status was changed from 0 to 1 and vice versa. BMI was investigated by increasing or reducing BMI by 10%. For all scenarios, the original and counterfactual inputs were entered into the pipeline and the model predicted the final PNT score for both scenarios. The difference in the predicted PNT score was calculated as the estimated impact of each counterfactual change. This was calculated individually for each health factor, and as a combined factor given that participants who had hypertension were more likely to also have diabetes or a high BMI. See Figure 3 for a comparison of data entered into the main model and the counterfactual models.

**Figure 3.**
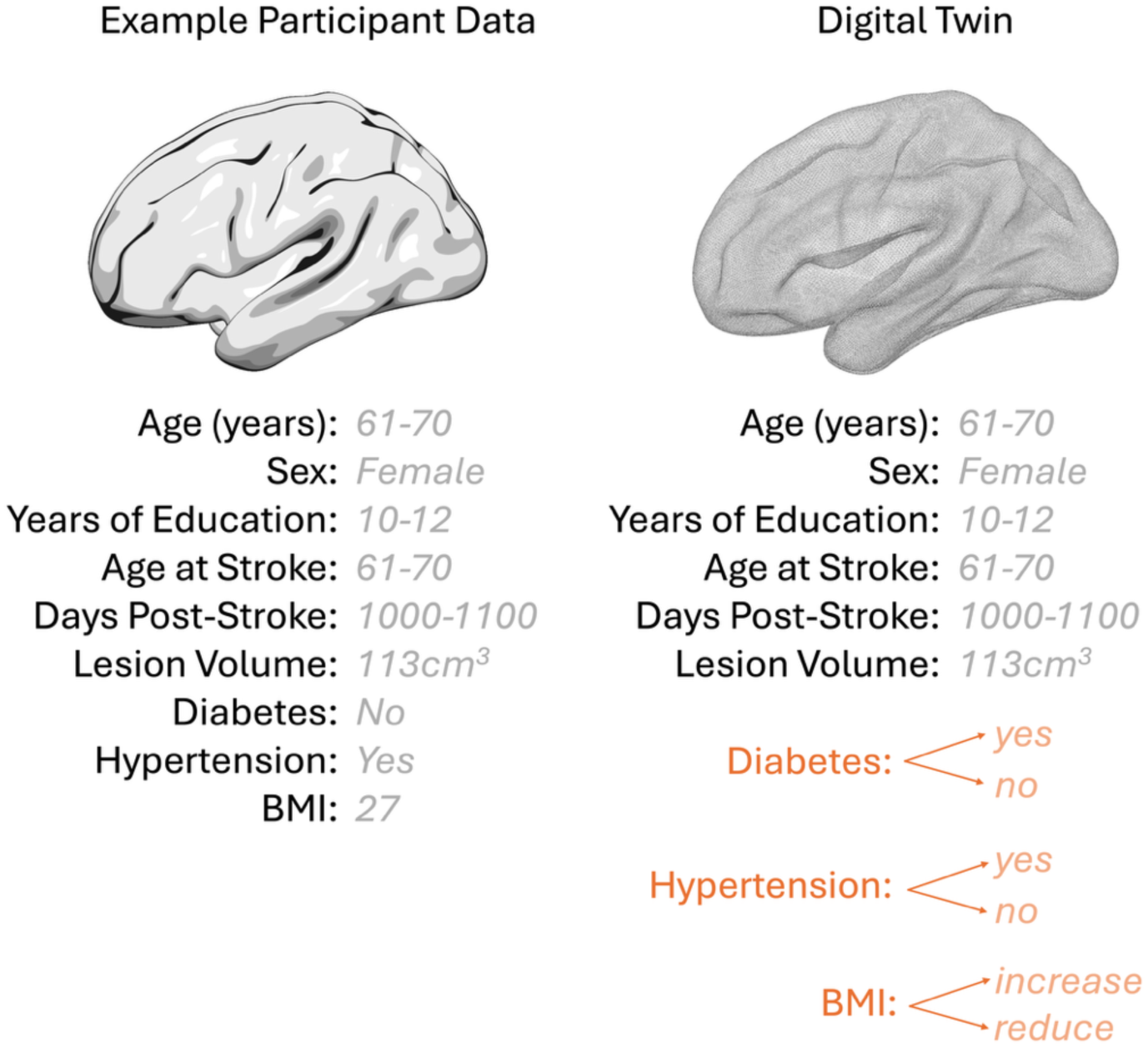
A comparison between the data entered into the main model and the counterfactual model.

To understand the impact of these counterfactual changes, we compared the predicted difference in PNT score with the average change in PNT score across the POLAR treatments.

## Results

Participants had a mean age of 60.64 years (SD = 10.87 years) and approximately 41% were female and 59% were male. See Table 1 for a full breakdown of participant characteristics, Figure 4 for overall participant information and Figure 5 for an example of the multimodal neuroimaging data for one participant.

**Table 1.** Participant characteristics (N = 106)

|  | Mean or % | SD or N |
| --- | --- | --- |
| **Demographic Variables** |  |  |
| Sex, Female | 40.57% | 43 |
| Chronological Age, Years | 60.64 | 10.87 |
| Education, Years | 15.33 | 2.27 |
| **Stroke Variables** |  |  |
| Age at Stroke, Years | 57.10 | 11.73 |
| Lesion Volume, cm<sup>3</sup> | 12.35 | 11.73 |
| Time Post-Stroke, Days | 1459.58 | 1516.71 |
| Stroke Type |  |  |
| Hemorrhagic | 27.36% | 29 |
| Ischemic | 64.15% | 68 |
| Other* | 8.49% | 9 |
| **Health Variables** |  |  |
| Diabetes | 22.64% | 24 |
| Hypertension | 59.06% | 52 |
| BMI | 27.21 | 4.97 |
| **Aphasia** |  |  |
| PNT | 59.69 | 22.85 |
| WAB AQ | 79.14 | 60.98 |
| Aphasia Type |  |  |
| Anomia | 28.30% | 30 |
| Broca's | 45.28% | 48 |
| Conduction | 15.09% | 16 |
| Global | 3.77% | 4 |
| Transcortical Motor | 0.94% | 1 |
| Wernicke's | 6.60% | 7 |

WAB-R AQ = Western Aphasia Battery-revised Aphasia Quotient, PNT = Philadelphia
\*Other refers to an instance where it is unclear if the stroke was hemorrhagic or ischemic.

**Figure 4.**
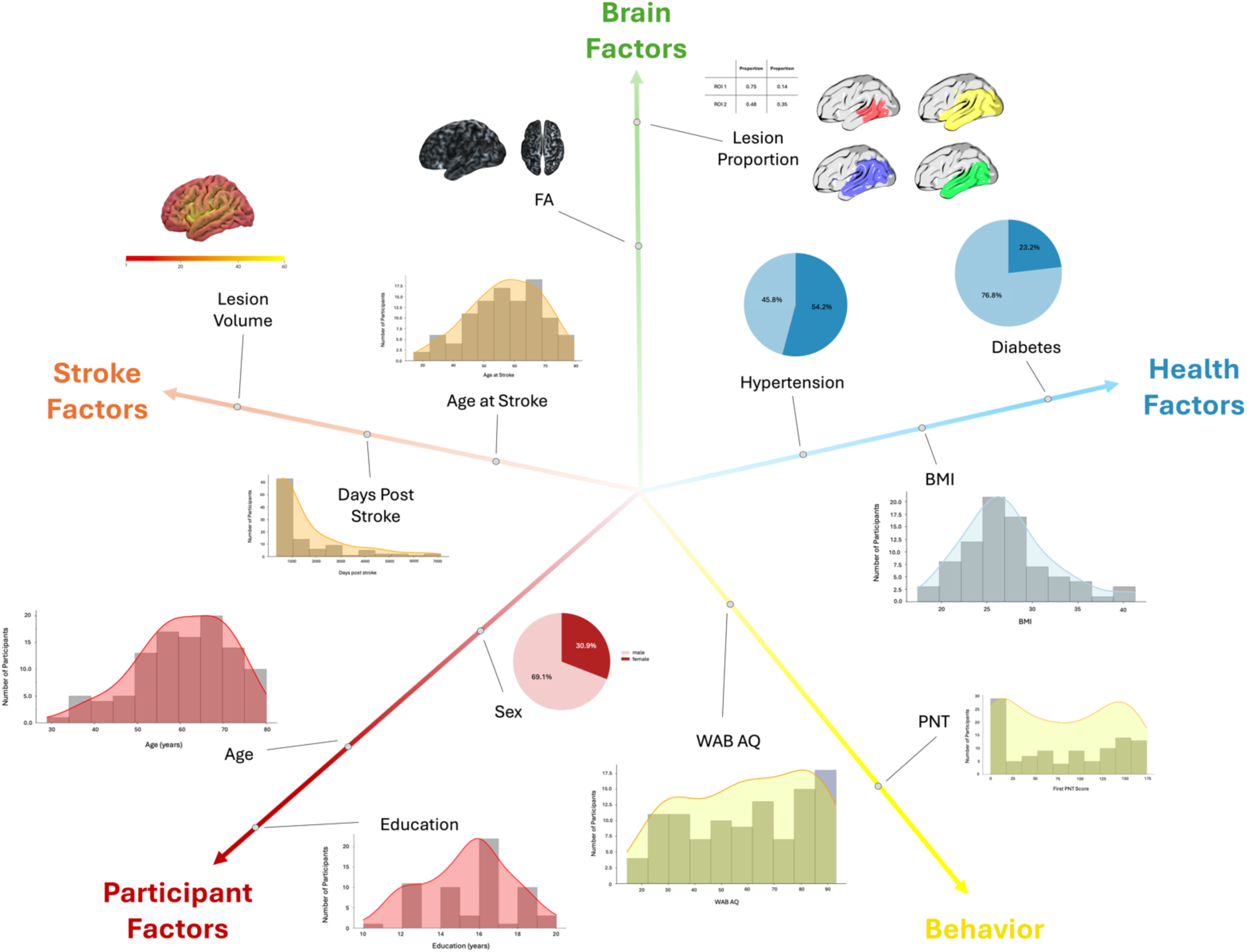
Factors included in the modeling framework, organized into five domains: participant factors (red; e.g., age, sex, education), stroke factors (orange; e.g., lesion volume, days post-stroke), brain factors (green; e.g., lesion proportion, hemispheric FA), health factors (blue; e.g., hypertension, diabetes, BMI), and behavioral performance (yellow; WAB AQ and PNT). Histograms and diagrams show the distribution and anatomical representation of each variable included in the model.

**Figure 5.**
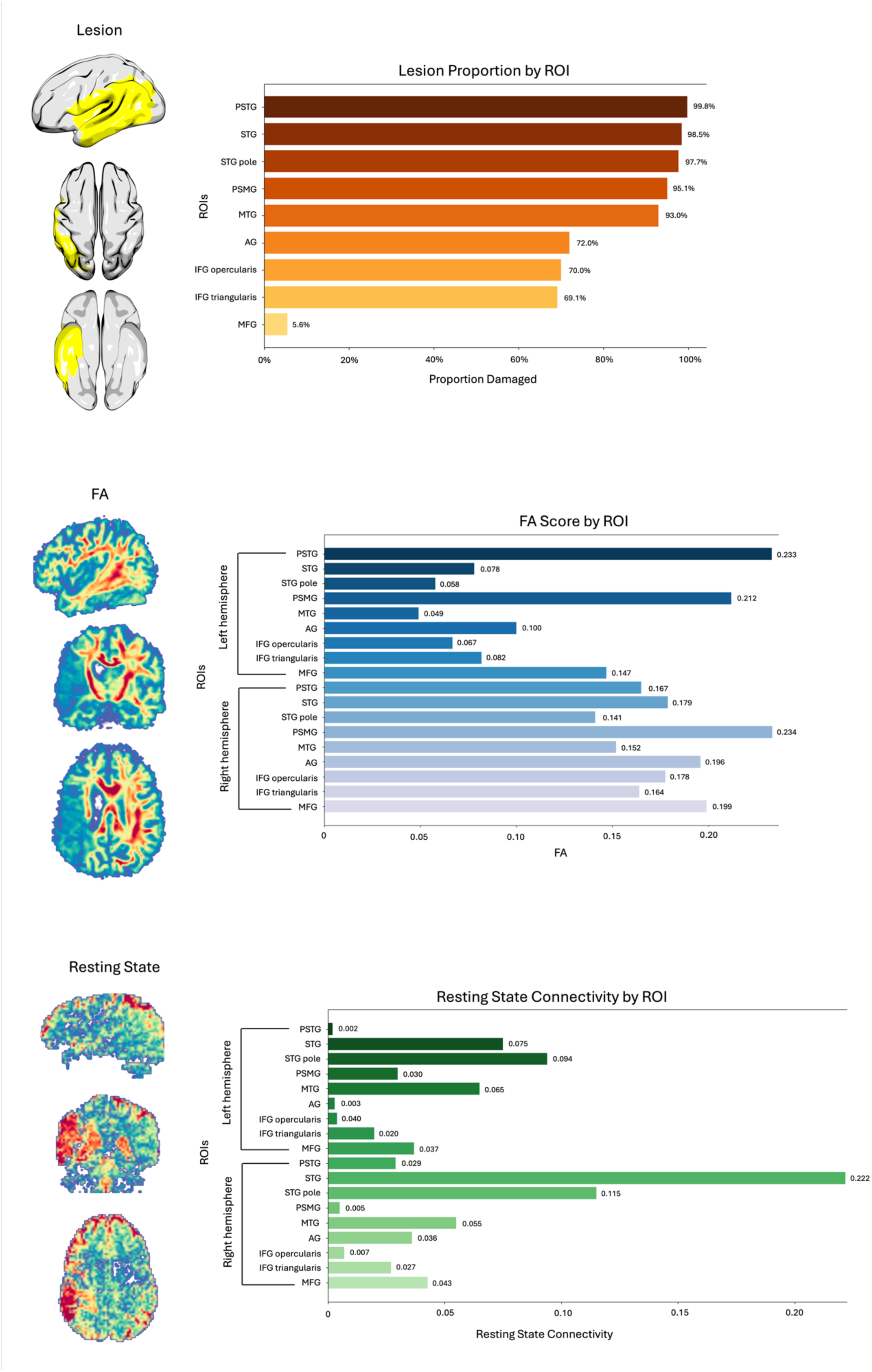
Multimodal neuroimaging data which went into the models for an individual participant. At the top is the lesion data, the middle shows the FA data and the bottom shows the resting state connectivity data.

When predicting the final PNT score for each participant, the model explained 58.48% of the variance (R^2^ = 0.5848). The factors that most contributed to the model were the previous PNTs (weight: 0.096), age (weight: -0.039), white matter integrity in right MTG (weight: 0.030), lesion load in PSMG (weight: -0.028), white matter integrity in right STG pole (weight: 0.028), days post-stroke (weight: 0.027), white matter integrity in right STG (weight: 0.027), white matter integrity in right PSMG (weight: 0.022), white matter integrity in left PSTG (weight: 0.022), lesion load of left AG (weight: -0.021). See Figure 6 for a visualization of these top factors and a breakdown of the contribution of all factors included in the model and Figure 7 for a brain map of the contributions of lesion load in language-related regions.

**Figure 6.**
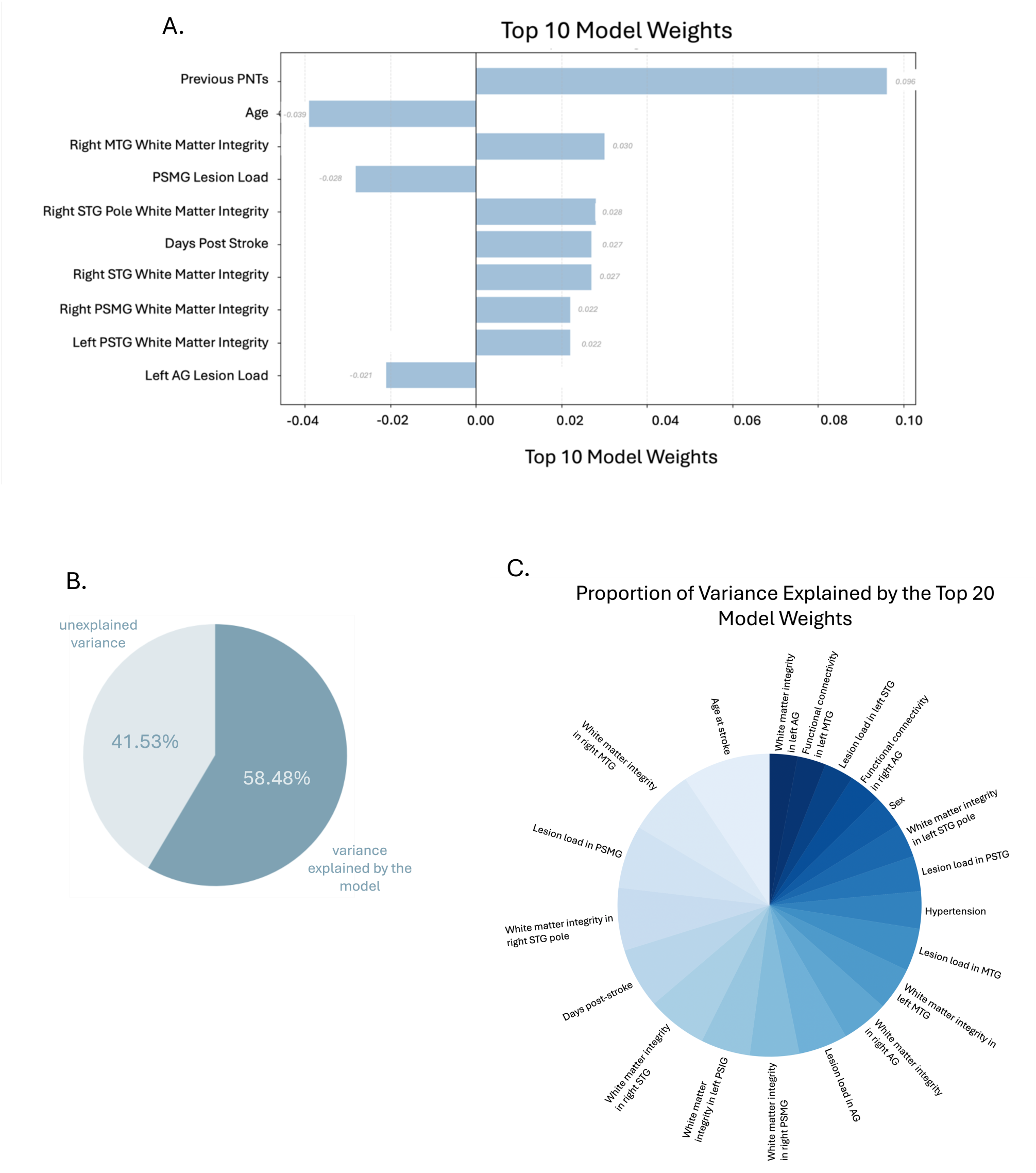
A shows the ten highest-weighted predictors in the main model, reflecting their relative influence on predicted PNT scores. B illustrates the overall variance in PNT performance explained by the model. C shows how the explained variance is apportioned across the top 20 individual predictors, demonstrating the relative importance of each factor within the digital twin model. Note that we have excluded previous behavioral performance from the pie chart to better illustrate the other contributing factors.

**Figure 7.**
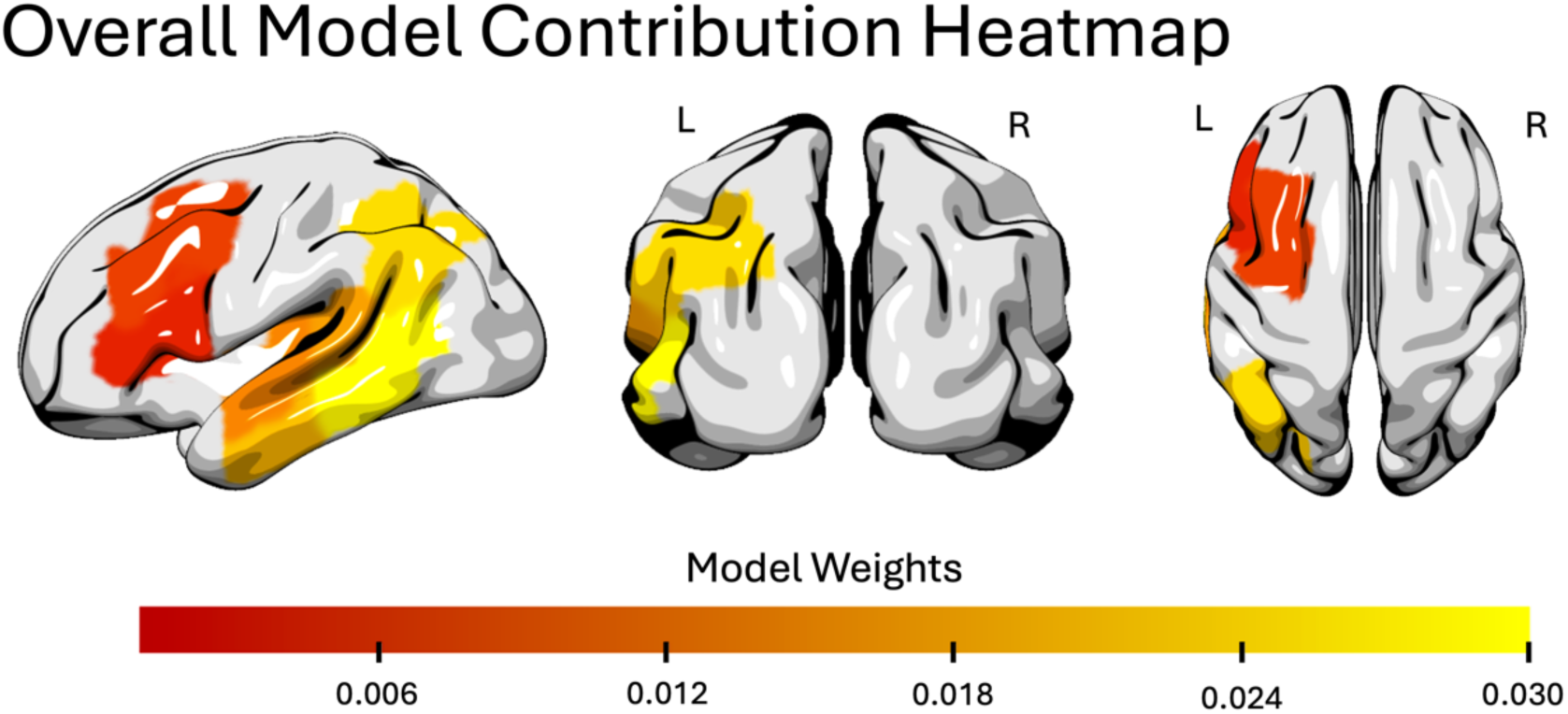
Heatmap illustrating the contribution of lesion load within left-hemisphere language-related regions to the digital twin model. Highlighted ROIs include AG, IFG triangularis, IFG opercularis, MFG, MTG, PSTG, PSMG, STG, and the STG pole. This visualization isolates lesion-derived contributions; hemispheric diHusion and resting-state connectivity measures were incorporated in the model but are not depicted here.

### Counterfactual Changes

The counterfactual simulations used in the model allow estimation of the contribution of each health factor by simulating scenarios in which the health of an individual or a set of individuals differs. Three health factors were tested: diabetes, hypertension, and BMI. For diabetes, changing the status of individuals who did not have diabetes to having diabetes was associated with an average 0.16 reduction in PNT score, whereas changing the status of those with diabetes to without diabetes was associated with a

0.09 increase in the PNT score. Similarly, changing hypertension status from no hypertension to hypertension was associated with a 0.38 point reduction in PNT score, while changing hypertension status from having hypertension to not having hypertension was associated with a 0.72 point increase on the PNT. Finally, increasing BMI by 10% was associated with a 1.48 point reduction in PNT, while reducing BMI was associated with a 1.48 point increase on the PNT. See Figure 8 for a visualization of these differences in predicted scores.

**Figure 8.**
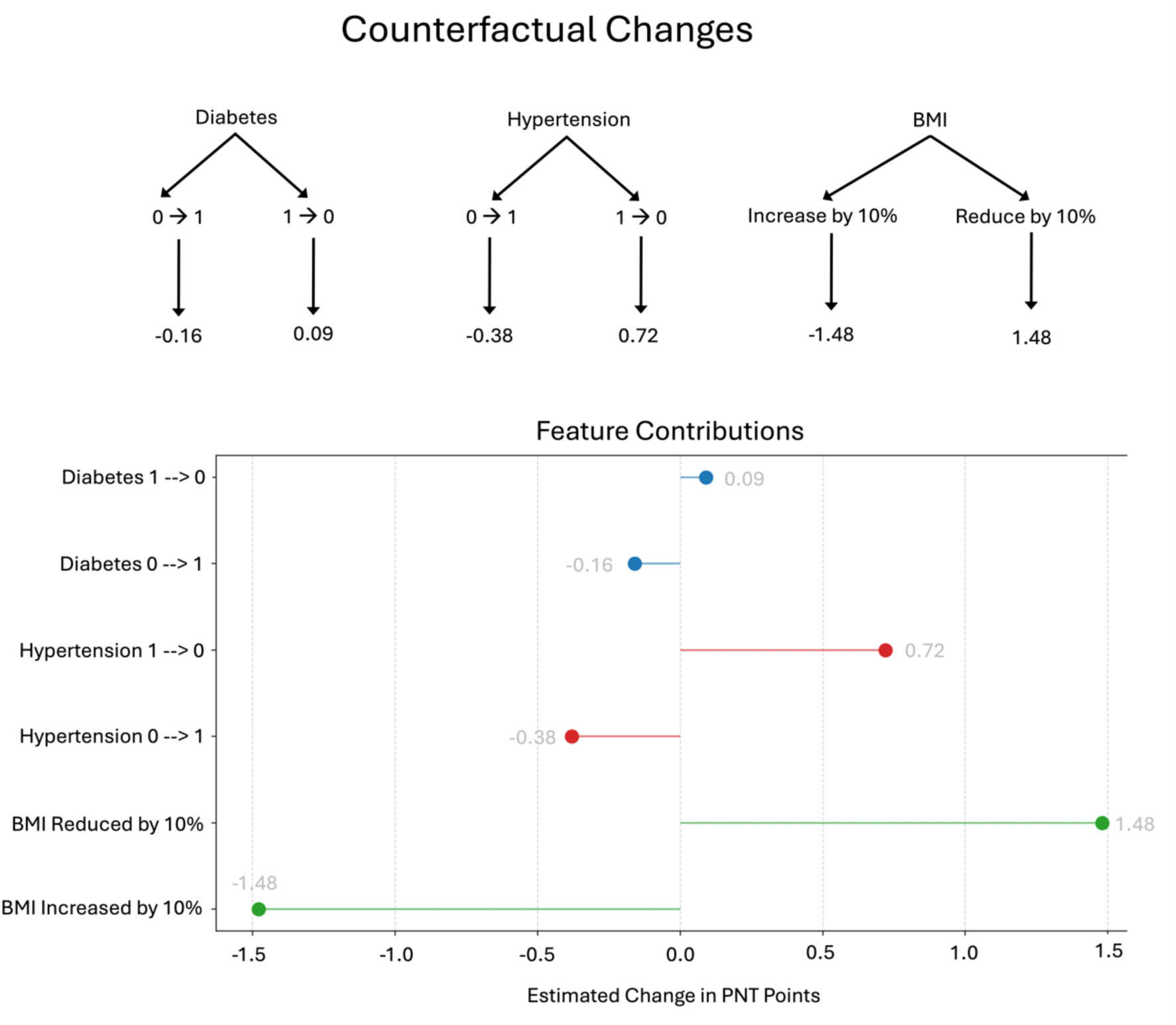
Outline of the counterfactual changes used in the model with the estimated change in PNT points after therapy (top) and a graph to display the estimated changes in PNT points after therapy (bottom).

Given that individuals who have one cardiovascular risk factor typically have more (i.e., people with diabetes are more likely to have hypertension and a high BMI), we also explored an omnibus model which incorporated changing both diabetic and hypertension status and increasing/decreasing BMI. If all three cardiovascular risk factors were changed (0 **→** 1 or 1 **→** 0) the average predicted change was a 2.02 point decrease or a 2.29 point increase in PNT score respectively. The largest estimated change in PNT score was 8.08 point and the lowest was 0 points.

The average change in PNT score across participants was approximately 8 points (exact average: 7.92, SD = 16.11). Therefore, diabetic status explained 2% of the variance in treatment gains, hypertensive status explained 4.75%, and increasing BMI explained 18.5%. Together, the health factors explained 25.25% of variance in treatment gains.

## Discussion

Digital twin models are increasingly popular tools in fields such as city planning and aerospace research, and they are an emerging concept in healthcare. Typically, digital twin models in medicine can span from high-fidelity biophysical simulations to pragmatic, data-driven clinical models. Because the full physiological complexity of post-stroke recovery involves many dynamic processes, any clinically useful digital-twin framework must begin with a restricted set of core features. The present study represents a foundational first step: a basic digital-twin model built from behavioural, demographic, health, and MRI-derived measures that are routinely available in clinical research. Although necessarily simplified, this approach establishes a translationally viable basic digital twin model that can perform patient-specific prediction and counterfactual reasoning. Importantly, the framework aligns with the emerging use of digital twins in clinical trials, where they are increasingly leveraged to simulate individual treatment responses, stratify participants, and quantify the potential impact of modifiable risk factors. By situating our model within this broader methodological landscape, we demonstrate how even a basic digital twin—constructed from real-world longitudinal stroke data—can begin to illuminate mechanisms of recovery using clinical trial data and can support personalised rehabilitation strategies.

In this study, we tested a digital twin model that predicts naming scores from multimodal data in chronic aphasia and estimates the potential impact of modifiable health factors on recovery trajectories through counterfactual simulation. The digital twin model could significantly predict approximately 60% of the variance in naming (PNT scores - R^2^ = 0.5848). By changing the status of health factors, we were able to explore how each factor influenced naming scores. We found that removing diabetes or hypertension from the digital twin positively impacted the estimated PNT score, albeit by a small margin. Conversely, increasing individual BMI by 10% was associated with a slightly larger (1.48 point) decrease in estimated PNT. Although these estimations are modest, the average change in PNT score across treatment was 8 points, therefore together these health factors explained 25% of the variance in treatment gains. Overall, this study demonstrates the potential utility of digital twin-modeling of aphasia recovery in the context of language treatment in the chronic stage. It also underscores the importance of accounting for health, demographic and physiological factors into account in cognitive recovery after strokes. With potential advances in physiological monitoring, the precision and clinical value of digital twin modeling in post-stroke cognitive recovery are likely to increase substantially.

### Main Prediction Model

The digital twin model captured a substantial portion of the variance in the most recent PNT score (R² = 0.58), despite the heterogeneity of stroke presentations and recovery pathways. This performance illustrates the value of integrating multimodal data (lesion characteristics, brain integrity, health factors, and demographics) within a single adaptive framework. Because the model updates with new information, it comprises a patient-specific and evolving prediction system rather than a static, group-derived estimate. This flexibility is crucial in stroke recovery, where clinical trajectories differ widely across individuals.

Previous naming performance emerged as the strongest predictor in the model, consistent with prior work demonstrating that pre-treatment PNT scores, and more generally, baseline aphasia severity, are robust indicators of subsequent therapy response and long-term language outcomes ^44–46^. Although baseline performance is partly influenced by structural damage, it is not reducible to lesion characteristics alone, underscoring its complementary role as a global measure of functional impairment. Lesion-related variables also made substantial contributions to prediction accuracy, particularly lesion burden within language-relevant regions such as the posterior supramarginal gyrus (PSMG), angular gyrus (AG), and middle temporal gyrus (MTG). These findings align with extensive evidence that structural preservation of core language regions supports better recovery trajectories ^4,47–50^, and reinforces the value of incorporating both behavioral and anatomical measures when modeling post-stroke

Other important factors were age and days post-stroke. Age-related influence on cognition in the general population is well documented ^51–55^, as assessed by tests such as the MoCA^56,57^ or the Mini Mental State Exam (MMSE)^56,58^, but the relationship between age and language-specific aspects of cognition is less clear. Some studies suggest that there is an age-related decline in name retrieval, but no corresponding decline in the retrieval of object and action-related words^59^. Similarly, aged-related changes in language production appear more notable than comprehension^55^. Our results support the notion of language production deficits with older age, but future studies could further investigate this more using comprehension-focused tasks. Although core language regions are thought to be relatively preserved in healthy aging^60^, it may be that some specific aspects of age-related cognitive decline may be associated with reductions in the structural integrity of the brain. It may also be that aging is associated with an increased incidence of other brain-health related problems, including hypertension or presence of WMHs, which could, in turn, influence behavior.

White matter integrity in language-relevant regions in the right hemisphere were also significant predictors in the overall model which suggests that the integrity of non-lesioned brain tissue may be an essential factor for post-stroke recovery. Small vessel disease is a biomarker of (structural) brain health. Post-stroke recovery is likely dependent on longer-term cerebrovascular health, which can be measured through white matter hyperintensities (WMHs)^21,61^, a marker of small vessel disease. WMHs indicate damage to white matter tracts across the brain, notably long-range association tracts which are important for complex behavior such as language^62–67^. For example, Wilmskoetter et al. demonstrated that small vessel brain disease post-stroke affects chronic aphasia severity through a change in the proportions of long- and short-range fibers^67^. Our results do not provide an explanation for the association between white matter integrity and PNT score, however they do bolster the idea that intact brain regions away from the lesion may be important predictors of recovery trajectories.

### Counterfactual Simulations

In this study, counterfactual simulations were important for modeling the impact of different health factors that static models cannot model. To explore the potential impact of hypertension on language recovery, we conducted counterfactual simulations in which participants’ hypertension status was modified. Specifically, we compared model predictions when hypertension was present versus absent, holding all other variables constant. The model consistently predicted higher PNT scores when hypertension was removed and lower PNT scores when hypertension was added. These bidirectional results, while modest, suggest that the presence of hypertension, even when controlling for lesion burden and other demographic factors, independently influences predicted recovery outcomes. While the model does not establish causality, these findings align with prior research linking hypertension to reduced cognition and overall brain health^19,68–71^ and suggest that hypertension may play a role in aphasia recovery. The ability to simulate individual- and group-level outcomes across different hypertension states could help to identify individuals who may benefit from more aggressive cardiovascular management and may offer interpretable predictions that could eventually inform personalized rehabilitation strategies.

Similarly, we also evaluated the impact of diabetes through counterfactual simulation and found the same bidirectional results. While the absolute differences were smaller than those observed for hypertension, the directionality of the effects remained consistent, suggesting that diabetes is also a negative predictor of language recovery, albeit with a more modest effect size. From a clinical perspective, these results reinforce the importance of vascular health in neurorehabilitation. Diabetes is associated with microvascular dysfunction, neuroinflammation, and impaired white matter integrity, all of which may compromise recovery potential following stroke. Although the observed effects were modest, the ability to simulate outcomes under different metabolic health conditions may help identify patients at greater risk for poor recovery. These findings further demonstrate how digital twin models can be used not only for prediction, but for exploring the impact of modifiable health factors on individual recovery trajectories.

In addition, we explored the impact of BMI on language recovery by increasing or decreasing BMI across all individuals. This resulted in a larger estimated difference than those observed for hypertension or diabetes. This effect was consistent across participants, suggesting that BMI plays a more prominent role in shaping predicted recovery outcomes in this sample. Given that BMI is a continuous variable, even modest reductions had measurable effects on predicted PNT scores, highlighting its potential importance as a modifiable health factor. Clinically, elevated BMI is associated with systemic inflammation, reduced cerebrovascular health, and structural brain changes which may influence recovery following a stroke. The digital twin’s sensitivity to BMI suggests that optimizing metabolic health could enhance the potential for language recovery. Importantly, the model allows simulation of potential benefits of BMI reduction at an individual level, offering a tool for personalized rehabilitation planning. These findings support growing evidence that metabolic and cardiovascular risk factors should be considered not just in stroke prevention, but also in post-stroke recovery interventions.

Although the effects of hypertension, diabetes, and BMI were modest when examined independently, their collective influence can amount to a significant influence on aphasia symptomatology and recovery, particularly when placed in the context of treatment gains within this clinical trial. Post-stroke recovery reflects the cumulative impact of multiple biological vulnerabilities rather than the effect of any single factor in isolation. Even small deviations in vascular or metabolic health may interact with each other—and with lesion characteristics—to constrain recovery potential in ways that are difficult to appreciate from unidimensional models. The digital twin framework enables simultaneous quantification of these influences, offering a more realistic representation of the interconnected factors that shape aphasia outcomes. By integrating incremental contributions across domains, the model highlights how the totality of an individual’s health profile can meaningfully alter their predicted trajectory. This accumulation of modest but converging effects underscores both the complexity of aphasia recovery and the value of approaches capable of capturing this complexity in a quantitative, individualized, and interpretable manner.

### Limitations

This study has several limitations. First, hypertension and diabetes were coded as binary variables, which restricted counterfactual simulations to flipping each condition from present to absent (or vice versa), rather than allowing finer-grained exploration of disease severity or physiological burden. Second, health measures were collected at a single time point, preventing us from incorporating temporal changes in metabolic or cardiovascular status over the study period. These constraints likely underestimate the true influence of health trajectories on recovery. Future studies would benefit from more detailed and continuously measured health data, enabling incremental counterfactual manipulations and broader participant inclusion, as was done with BMI, treated as a continuous variable, here.

An additional limitation is that we only change BMI by 10% up or down, and assume that lowering BMI is beneficial and increasing BMI is associated with poorer overall health. However, it is important to note that a 10% change in BMI is typically 1-3 points so these changes will have only affected individuals who are on the border of BMI categories.

### Conclusions

This study demonstrates the feasibility and clinical potential of applying a digital twin framework to chronic post-stroke aphasia. The digital twin model of chronic post-stroke aphasia successfully predicted more than half of the variance in naming performance during language treatment. By incorporating multimodal neuroimaging, demographic, and health-related data, the model captured the majority of the variance in individual recovery trajectories. Through counterfactual simulation, we demonstrated that modifiable health factors, including hypertension, diabetes, and BMI, exert measurable, bidirectional influences on predicted treatment outcomes, underscoring the role of systemic health in shaping language recovery. Although the individual effects of hypertension, diabetes, or BMI were modest in magnitude, their cumulative influence on treatment gains illustrates how multiple small biological contributors can add up to shape meaningful differences in language outcomes.

More broadly, these findings illustrate the potential value of digital twin models for aphasia treatment, particularly as a tool to integrate diverse biological factors and generate individualized, dynamically updated predictions. As physiological, metabolic, and behavioral data become more routinely collected in clinical settings, digital twins may offer increasingly accurate representations of patient-specific trajectories. Importantly, while the present model was applied in the context of treatment-related recovery in the chronic stage, the same framework could be extended to predict spontaneous recovery at earlier time points, including the acute and subacute phases. With additional longitudinal data, digital twin models may provide a unified platform for forecasting outcomes, potentially guiding the type and timing of therapeutic decisions, and identifying modifiable health features that could enhance recovery across the full course of post-stroke rehabilitation.

## Data Availability

The data that support the findings of this study are available from the corresponding author upon reasonable request. Ethics restrict our ability to share identifiable patient data, including behavioral, health data, or imaging data. Therefore, a reasonable request in this instance refers to a request that does not include identifiable data.

